# Classification of Pediatric Dental Diseases from Panoramic Radiographs using Natural Language Transformer and Deep Learning Models

**DOI:** 10.1101/2025.01.30.25321418

**Authors:** Tuan D. Pham

## Abstract

Accurate classification of pediatric dental diseases from panoramic radiographs is crucial for early diagnosis and treatment planning. This study explores a text-based approach using a natural language transformer to generate textual descriptions of radiographs, which are then classified using deep learning models. Three models were evaluated: a one-dimensional convolutional neural network (1D-CNN), a long short-term memory (LSTM) network, and a pretrained bidirectional encoder representations from transformer (BERT) model for binary disease classification. Results showed that BERT achieved 77% accuracy, excelling in detecting periapical infections but struggling with caries identification. The 1D-CNN outperformed BERT with 84% accuracy, providing a more balanced classification, while the LSTM model achieved only 57% accuracy. Both 1D-CNN and BERT surpassed three pretrained CNN models trained directly on panoramic radiographs, indicating that text-based classification is a viable alternative to traditional image-based methods. These findings highlight the potential of language-based models for radiographic interpretation while underscoring challenges in generalizability. Future research should refine text generation, develop hybrid models integrating textual and image-based features, and validate performance on larger datasets to enhance clinical applicability.

## 1 Introduction

Accurate and early diagnosis of pediatric dental diseases is essential for effective treatment planning and the prevention of long-term complications [1, 2, 3, 4, 5]. Panoramic radiographs are widely used in dentistry to assess conditions such as caries and periapical infections. Traditionally, diagnosis relies on manual interpretation by dental professionals, a process that can be time-consuming and subject to interobserver variability. To enhance diagnostic efficiency and accuracy, artificial intelligence (AI) has emerged as a promising tool in dental imaging, particularly through deep learning models trained for automated disease classification [6, 7, 8, 9].

Recent advances in deep learning have enabled AI models to assist in detecting and diagnosing a range of pediatric dental conditions, an area that has historically received less attention than adult dental diagnostics. Kaya et al. [10] introduced a deep learning model specifically designed to detect permanent tooth germs in pediatric panoramic radiographs, addressing a gap in prior research, which has primarily focused on adult radiographs. Similarly, Ong et al. [11] developed a fully automated deep learning framework for assessing dental development stages in pediatric radiographs, further demonstrating AI’s potential to improve diagnostic accuracy and clinical decision-making.

The systematic review by Khanagar et al. [12] highlights the transformative impact of AI in pediatric dentistry, particularly in early disease detection, which is critical for timely intervention and treatment. Their review examined various AI models applied in diagnosing conditions such as dental caries and periodontal diseases, reinforcing the potential of machine learning in pediatric oral healthcare. Beyond disease detection, AI also plays a role in treatment planning and patient management. La Rosa et al. [13] explored how AI algorithms can identify early signs of dental pathologies and improve orthodontic diagnoses through automated image analysis. This aligns with Mahajan et al. [14], who conducted a systematic review on the effectiveness of AI in pediatric dentistry, concluding that AI-based diagnostic tools have demonstrated promising results in clinical applications.

Despite these advancements, the implementation of AI in pediatric dentistry faces several challenges, particularly concerning data quality, dataset size, and model generalizability. AI models require large and diverse datasets to achieve robust performance, yet pediatric dental datasets are often limited, leading to potential biases and decreased model reliability. Hsieh [15] emphasized that while AI holds great promise for comprehensive dental disease classification, challenges such as class imbalance and the rarity of certain conditions can hinder model training and diagnostic accuracy. These limitations underscore the need for ongoing research to refine AI methodologies, improve data preprocessing techniques, and develop strategies for overcoming data scarcity.

The integration of AI into pediatric dental disease classification from panoramic radiographs is a rapidly evolving field with significant potential to enhance diagnostic accuracy, reduce human error, and improve patient outcomes. However, to fully realize its benefits, further research is needed to address challenges related to data availability, annotation consistency, and model validation across diverse populations. While AI-driven image-based classification using convolutional neural networks (CNNs) has demonstrated success, such approaches often require extensive preprocessing as well as large datasets to achieve high performance. Hybrid models that integrate text descriptions from images and deep learning for classification could provide a promising direction for improving accuracy, interpretability, and scalability in pediatric dental diagnostics.

Large language models (LLMs), particularly ChatGPT, have emerged as transformative tools in medical and dental applications, leveraging advanced natural language processing (NLP) capabilities to enhance various aspects of healthcare delivery. These models are designed to understand and generate human-like text, making them valuable for a range of tasks, including patient communication, clinical decision support, and medical education.

One of the primary applications of ChatGPT in healthcare is its ability to assist in clinical decision-making. A study by Giannakopoulos et al. [16] evaluated the performance of ChatGPT and other LLMs in supporting evidence-based dentistry, demonstrating that these models can effectively provide relevant information and aid practitioners in making informed decisions, ultimately improving patient care. Similarly, Huang et al. [17] highlighted ChatGPT’s potential in dental education, suggesting that it could serve as a valuable resource for students by offering instant access to information and facilitating interactive learning experiences.

In patient communication, LLMs like ChatGPT have shown promise in addressing inquiries and enhancing patient satisfaction. A survey conducted by Yong et al. [18] found that LLMs were effective in resolving patient complaints, emphasizing their ability to generate thoughtful and contextually appropriate responses. This capability is particularly valuable in fostering better patient-provider relationships and improving overall healthcare experiences.

The integration of LLMs in medical education has also been a focal point of recent research. Abd-Alrazaq et al. [19] explored how these models enhance medical students’ learning experiences by providing comprehensive explanations of complex medical concepts. Similarly, Wu et al. [20] evaluated ChatGPT’s performance on nursing licensure examinations, indicating that LLMs can serve as effective study aids, potentially improving educational outcomes.

Despite these advantages, the deployment of LLMs in healthcare is not without challenges. Lin et al. [21] noted that while ChatGPT offers valuable support in critical care medicine, it also has limitations, such as the potential to generate inaccurate or misleading information. This concern is echoed by Tiwari et al. [22], who conducted a systematic review on the implications of ChatGPT in public health dentistry, highlighting the need for rigorous assessment of risks, including biases and misinformation, before widespread adoption.

The motivation for this study stems from the need to enhance the automated classification of pediatric dental diseases from panoramic radiographs by leveraging both NLP and deep learning. Traditional image-based classification methods often require extensive preprocessing, high-resolution datasets, and expertise to interpret complex radiographic features. To overcome these challenges, this study explores an alternative approach by using ChatGPT to generate textual descriptions of the conditions depicted in panoramic radiographs. These textual representations serve as input for deep learning models, enabling disease classification through language-based analysis rather than direct image interpretation. This approach aims to improve diagnostic accuracy, enhance model interpretability, and reduce reliance on large-scale labeled radiographic datasets. By integrating AI-driven text generation with deep learning classification, this study seeks to demonstrate the feasibility of a text-based framework for dental disease diagnosis, offering a scalable and interpretable alternative to traditional radiographic analysis.

## 2 Methods

### 2.1 Panoramic radiographs

The children’s dental panoramic radiographs dataset [23], hosted on Figshare [24], is utilized in this study. The dataset, titled *Child Dental Disease Detection Dataset*, was originally designed with a structured division into “Train” and “Test” subsets, as described in [23]. Each subset contains both the original radiographic images and corresponding expert annotations. The training set consists of 70 images categorized into five dental diseases, with 14 images per category: caries (class 1), periapical infections (class 2), pulpitis (class 3), deep sulcus (class 4), and dental developmental abnormalities (class 5).

The annotation process involved six dental experts to ensure accuracy and reliability. Four experts independently annotated 25 randomly assigned anonymous images in the first round. Two additional experts then reviewed the labeled images to assess annotation accuracy. In cases of ambiguity, all six experts engaged in a consensus discussion, and the final labels were determined based on their unified decision. This multi-step validation process aimed to enhance the consistency and diagnostic precision of the dataset annotations.

For this study, only class 1 (caries) and class 2 (periapical infections) are utilized to maintain a balanced sample size for machine learning classification. The Test dataset includes 15 images for each of these two classes, making a total of 29 images (14 from the training set and 15 from the test set) available for classification. This selection ensures a more robust and statistically balanced dataset, improving the reliability of machine learning model evaluation.

### 2.2 LLM: ChatGPT

ChatGPT [25] is an advanced LLM based on OpenAI’s GPT architecture, designed to process and generate human-like text by leveraging deep learning techniques. Trained on vast datasets, including general knowledge, medical literature, and structured language patterns, ChatGPT can understand, summarize, and generate contextual responses across diverse domains. Its transformer-based architecture allows it to analyze complex relationships between words, enabling it to generate coherent and contextually relevant descriptions. This makes ChatGPT a valuable tool in medical AI applications, particularly for interpreting and summarizing clinical information.

Although ChatGPT does not directly analyze images, it can process structured input derived from automated or human-interpreted imaging analyses. By leveraging pre-existing medical knowledge, it can describe abnormalities such as bone loss, periapical radiolucencies, or carious lesions based on textual cues extracted from images. High-resolution panoramic radiographs contain detailed anatomical structures, but reduced-resolution images may lose some fine details. ChatGPT can compensate for this by generating structured descriptions that highlight key diagnostic features, ensuring that relevant clinical insights are preserved for downstream classification tasks.

A text-based approach provides an interpretable summary of radiographic findings, making AI-driven diagnostic models more accessible to clinicians. ChatGPT-generated descriptions allow for a structured, standardized representation of dental disease characteristics, improving transparency in AI-assisted diagnosis. Additionally, processing full-resolution radiographs with deep learning models requires substantial computational resources. By reducing image resolution and transforming key features into text, ChatGPT enables a more computationally efficient pipeline while retaining essential diagnostic information. By converting panoramic radiograph features into text, ChatGPT serves as a bridge between image interpretation and deep learning classification, offering an efficient, interpretable, and scalable approach to dental disease diagnosis.

### 2.3 LSTM

LSTM networks [26] are a type of recurrent neural network (RNN) designed to handle sequential data by addressing the issue of vanishing gradients, which commonly affects traditional RNNs. LSTMs are particularly effective in learning long-range dependencies, making them well-suited for tasks involving time series, NLP, and speech recognition.

Unlike standard RNNs, which struggle to retain information over long sequences, LSTMs use a specialized gating mechanism to regulate the flow of information. The network consists of three key gates: the forget gate, which decides what information to discard from the previous state; the input gate, which determines what new information to store; and the output gate, which controls what information is passed to the next time step. These gates allow LSTMs to selectively remember or forget information, enabling them to capture dependencies across long sequences while mitigating issues related to gradient decay.

The architecture of the LSTM model used in this study consists of a sequence input layer with an input size of 1, followed by a word embedding layer with an embedding dimension of 50. An LSTM layer with 100 hidden units was included, with the output mode set to “last” to process the entire sequence. A fully connected layer was added, with the number of units matching the number of classes in the data. A dropout layer with a rate of 0.2 was applied to prevent overfitting, followed by a softmax layer to classify the input into one of the two classes. Figure 1(a) shows the architecture of the LSTM.

**Figure 1:**
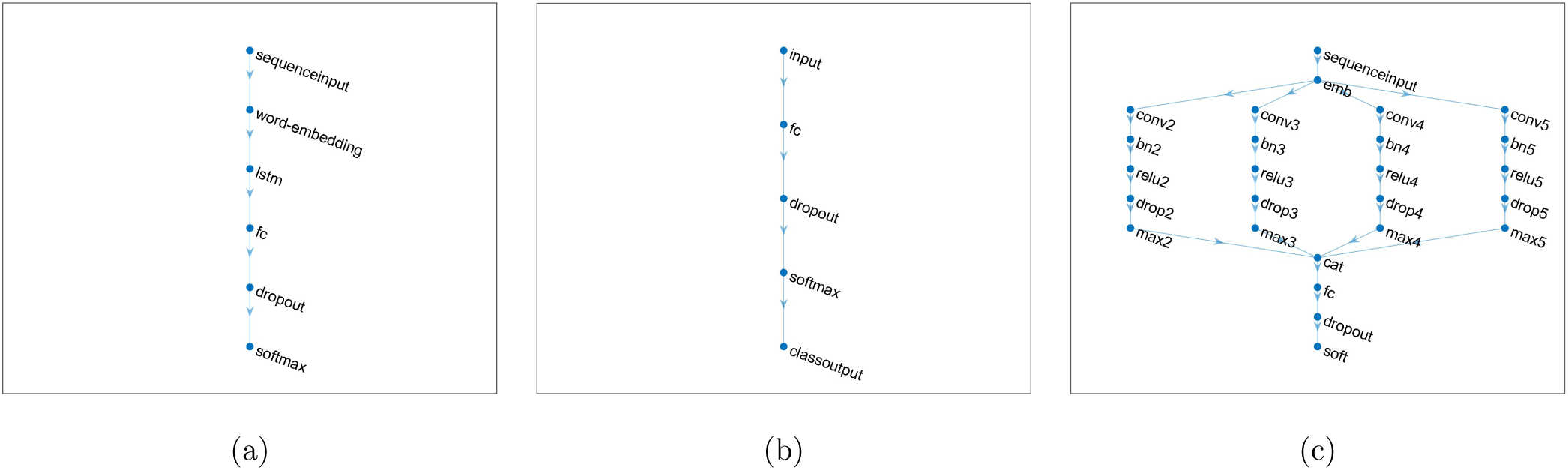
Network layers: LSTM (a), model trained with features extracted from BERT (b), and 1D-CNN (c).

For training, the Adam optimizer and cross-entropy loss were chosen, and the maximum number of epochs was set to 600. The batch size was specified as 128, and the model was evaluated using accuracy. Validation data was provided, and the best network based on validation performance was saved.

### 2.4 BERT

BERT [27] is a deep learning model, designed to process and understand natural language with high contextual awareness. Unlike traditional word embedding techniques that represent words independently, BERT captures deep bidirectional contextual relationships by considering both preceding and following words in a sentence. This ability makes it particularly powerful for tasks such as text classification, question answering, and language inference.

BERT is based on the transformer architecture, which relies on self-attention mechanisms to process entire sequences in parallel rather than sequentially, as in RNNs and LSTM networks. The model is pretrained on large text corpora using two key objectives: masked language modeling, where random words are masked, and the model learns to predict them, and next sentence prediction, where the model learns relationships between sentence pairs. This pretraining enables BERT to generalize well to various NLP tasks after fine-tuning on domain-specific datasets.

In this study, the BERT-Base model, consisting of 108.8 million learnable parameters, was used for classification. A tokenizer was employed to encode the text into sequences of integers, enabling the model to process the input effectively. Following tokenization, the data was partitioned into training and validation sets as previously described. To prepare the data for training, the BERT tokens were organized into mini-batches. This step ensures that the data is processed in manageable chunks, which is crucial for training large models like BERT. The BERT model was then used to transform the tokenized data into feature vectors by extracting embeddings, which served as input features for both the training and validation datasets.

Subsequently, a deep learning network was constructed for classification. The network architecture included a feature input layer, a fully connected layer to map the feature vectors to class labels, a dropout layer to reduce overfitting through regularization, and a softmax layer to output the class probabilities. This network was specifically designed to classify the BERT-derived feature vectors. Figure 1(b) shows the architecture of the network designed for classifying BERT-derived feature vectors, referred to as BERT for brevity.

The training process was configured with the following options: a mini-batch size of 128, the Adam optimizer, a maximum of 600 epochs, an initial learning rate of 0.0001, and without data shuffling to ensure consistent data order during training.

### 2.5 1D-CNN

For classification using a 1D-CNN [28], the network architecture was designed for text classification and begins with specifying the input size as 1, which corresponds to the channel dimension of the input integer sequence. The input data were then embedded using a word embedding layer with a dimension of 50.

The next step involved creating blocks of layers for different *n*-gram lengths, specifically 2, 3, 4, and 5. Each block consists of a 1D convolutional layer, batch normalization, a ReLU activation layer, a dropout layer with a rate of 0.2, and a global max pooling layer. For each block, 100 convolutional filters were used, and the size of the convolutional filter corresponds to the *n*-gram length. These blocks were connected to the word embedding layer, and their outputs were concatenated using a concatenation layer.

The final part of the architecture includes a fully connected layer, which outputs the class predictions, followed by a softmax layer for classification. The network was structured to handle various *n*-gram lengths by connecting each block to the word embedding layer and finally linking the pooling layers to the concatenation layer. Figure 1(c) shows the architecture of the 1D-CNN.

The network was trained using the Adam optimizer with a mini-batch size of 128. The model was optimized using cross-entropy loss, The training included validation using a separate validation dataset. The network with the lowest validation loss was saved.

### 2.6 Binary classification performance metrics

In this binary classification task, the objective is to distinguish between caries and periapical infections. The classification parameters are defined as follows: true positive (TP) refers to periapical infections correctly identified as periapical infections, while false positive (FP) denotes caries incorrectly classified as periapical infections. True negative (TN) represents caries accurately identified as caries, and false negative (FN) refers to periapical infections incorrectly classified as caries.

The performance of the classification model is evaluated using several key metrics. Accuracy (ACC) measures the proportion of correct predictions, including both true positives and true negatives, relative to the total number of predictions for the two classes. Sensitivity (SEN) reflects the model’s ability to correctly identify periapical infections, while specificity (SPE) assesses its ability to accurately classify caries. Precision (PRE) quantifies the proportion of cases predicted as periapical infections that actually belong to the class of periapical infection.

The F1 score is a metric that provides the harmonic mean of precision and sensitivity, balancing the trade-off between the two. It emphasizes the correct identification of periapical infections (true positives) while accounting for the impact of false positives and false negatives.

The area under the receiver operating characteristic curve (AUC) offers a comprehensive measure of the model’s ability to separate the two classes. It represents the relationship between sensitivity and specificity, with a higher AUC indicating better performance and greater robustness in distinguishing between caries and periapical infections.

Mathematical expressions for these performance metrics are provided in Table 1.

**Table 1:** Mathematical expressions for metrics evaluating the binary classification model.

| **Metric** | **Expression** |
| --- | --- |
| Accuracy (ACC) | $\frac{TP+TN}{TP+TN+FP+FN}$ |
| Sensitivity (SEN) | $\frac{TP}{TP+FN}$ |
| Specificity (SPE) | $\frac{TN}{TN+FP}$ |
| Precision (PRE) | $\frac{TP}{TP+FP}$ |
| F1 score | $2 \times \frac{PRE \times SEN}{PRE + SEN}$ |

## 3 Results

The resolution of the original panoramic radiographs was reduced 25 times. ChatGPT based on OpenAI’s GPT-4 architecture was then used to generate concise, approximately 12-word descriptions of potential bone abnormalities depicted in the reduced-resolution images. Figure 2 displays reduced-resolution panoramic radiographs showing caries and periapical infections in children, while Table 2 presents the ChatGPT-generated descriptions of the bone abnormalities corresponding to these radiographs.

**Figure 2:**
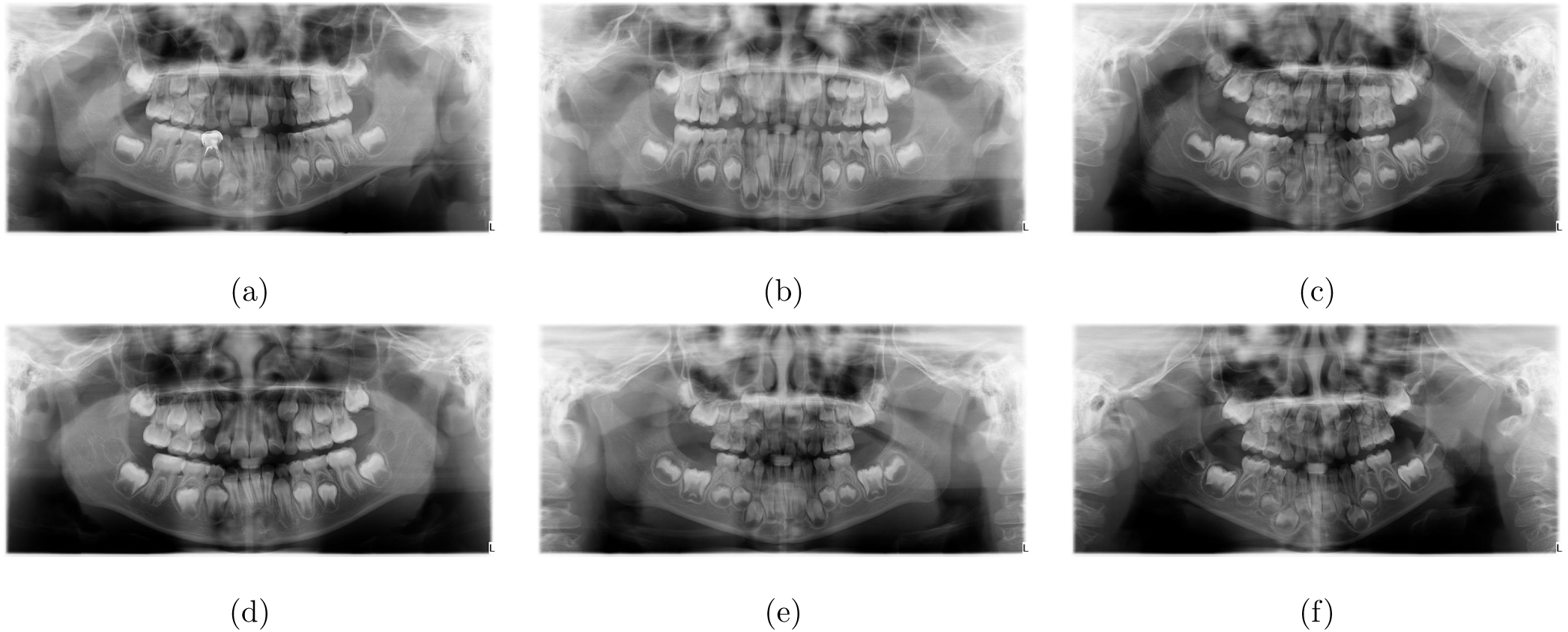
Reduced-resolution panoramic radiographs of pediatric dental diseases: caries or class 1 (a)-(c), and periapical infections or class 2 (d)-(f)

**Table 2:**
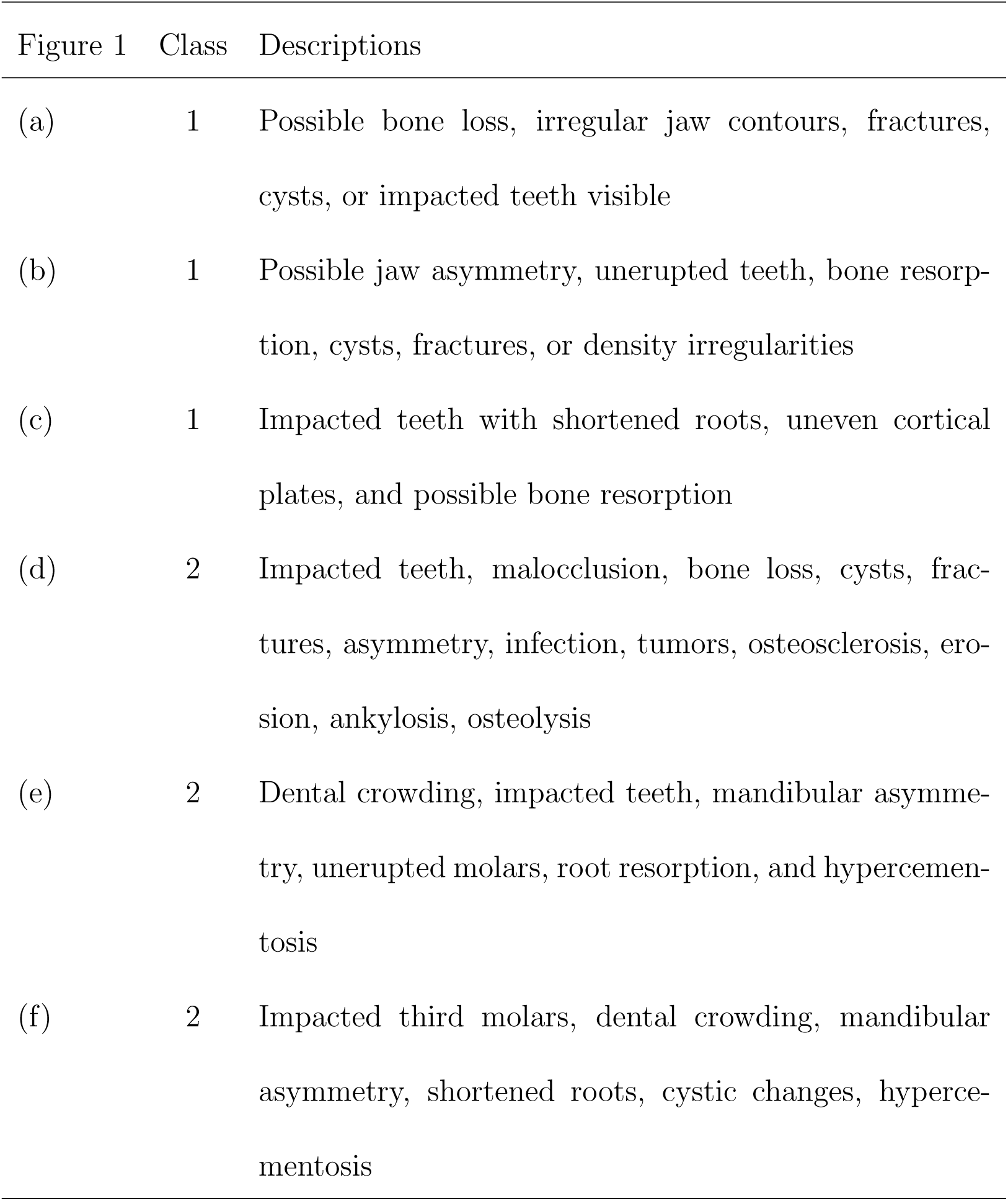
ChatGPT-based descriptions of pediatric dental diseases on panoramic radiographs as shown in Figure 2.

| Figure 1 | Class | Descriptions |
| --- | --- | --- |
| (a) | 1 | Possible bone loss, irregular jaw contours, fractures, cysts, or impacted teeth visible |
| (b) | 1 | Possible jaw asymmetry, unerupted teeth, bone resorption, cysts, fractures, or density irregularities |
| (c) | 1 | Impacted teeth with shortened roots, uneven cortical plates, and possible bone resorption |
| (d) | 2 | Impacted teeth, malocclusion, bone loss, cysts, fractures, asymmetry, infection, tumors, osteosclerosis, erosion, ankylosis, osteolysis |
| (e) | 2 | Dental crowding, impacted teeth, mandibular asymmetry, unerupted molars, root resorption, and hypercementosis |
| (f) | 2 | Impacted third molars, dental crowding, mandibular asymmetry, shortened roots, cystic changes, hypercementosis |

For the processed text data, each document is converted into a sequence of numeric indices. To achieve this, a word encoding function was used to create a word encoding, which maps words to numeric indices. The documents were then converted into sequences, ensuring that all sequences are of equal length. The sequences were padded and truncated to a target length, and the longest sequence length was selected for uniformity.

The text data were split into training and validation sets using a non-stratified holdout partition, where 90% of the data was allocated for training and the remaining 10% for validation. This partitioning process was repeated 10 times, and the average classification results were recorded. Figure 3 shows the word clouds of training and test data.

**Figure 3:**
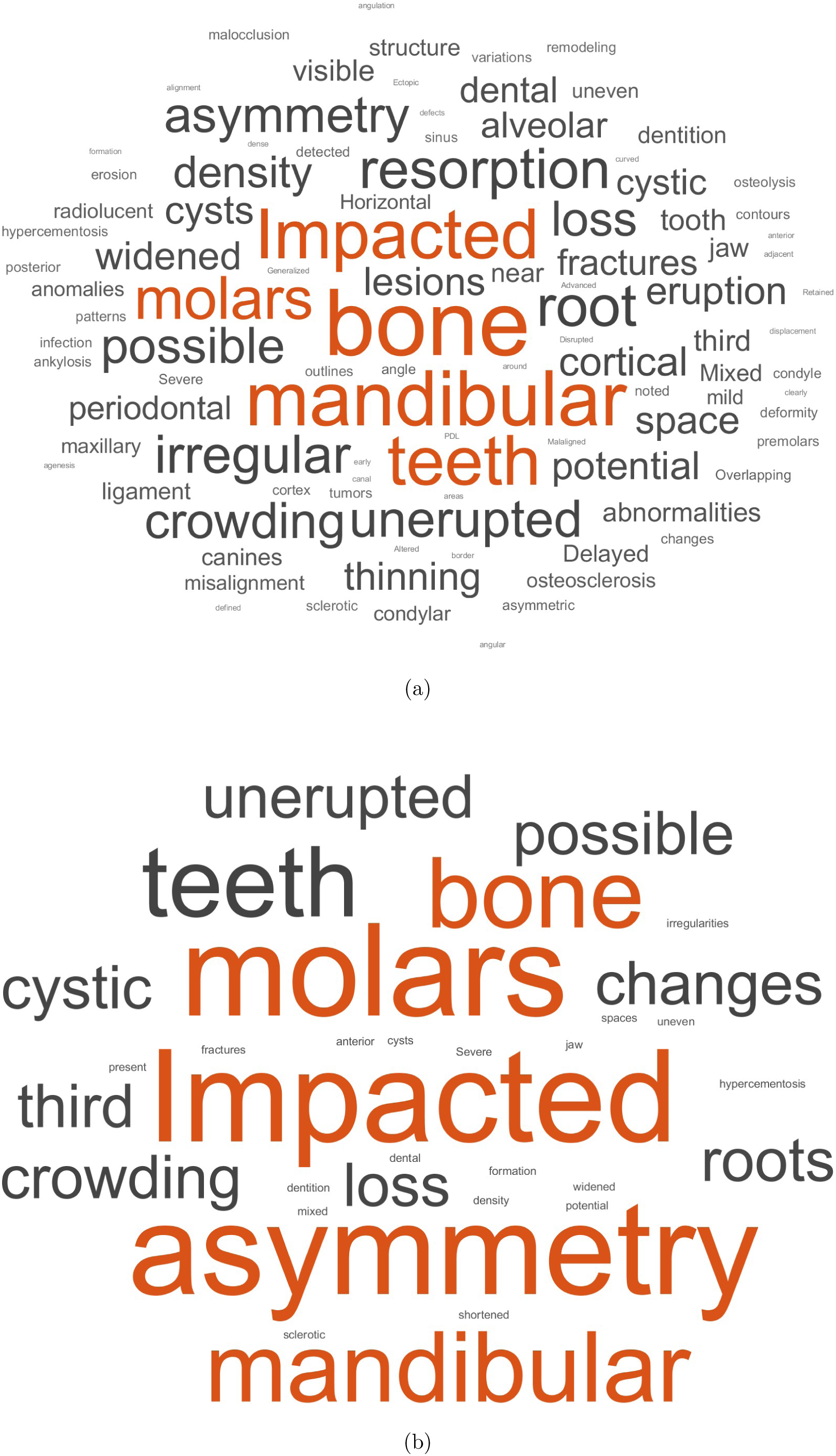
Word clouds of training (a) and test (b) data.

A preprocessing function is defined to prepare the text data for model input. This function performs several tasks: it tokenizes the text, converts it to lowercase, and removes punctuation. The training and validation text data are then processed using this function.

To compare the text-based with image-based classifiers, three pretrained CNNs that are SqueezeNet [29], GoogLeNet [30], and AlexNet [31] were also applied for the classification. The pretrained CNN models were trained and validated with reduced-resolution panoramic radiographs. Training options for the pretrained CNNs were specified as follows. The training process included data augmentation to enhance model generalization and prevent overfitting. Training images underwent random vertical flipping, translation of up to 30 pixels, and scaling variations between 90% and 110% to introduce variability and ensure the model learned good features rather than memorizing specific details. Validation images were resized without augmentation to maintain consistency during evaluation. The optimization method used is Adam, with a mini-batch size of 128 and a small initial learning rate of 0.0001, allowing stable and gradual updates. The models was trained for 100 epochs with data shuffling applied at every epoch to avoid learning biases. Validation accuracy was computed once per epoch,

Figure 4 illustrates the training and validation processes for the 1D-CNN, LSTM, and BERT models on a held-out partition. Figure 5 shows the training and validation processes for the three pretrained CNN models (SqueezeNet, GoogLeNet, and AlexNet) on a held-out partition. Table 3 presents the performance metrics obtained from the 1D-CNN, LSTM, BERT, and the three pretrained CNN models.

**Figure 4:**
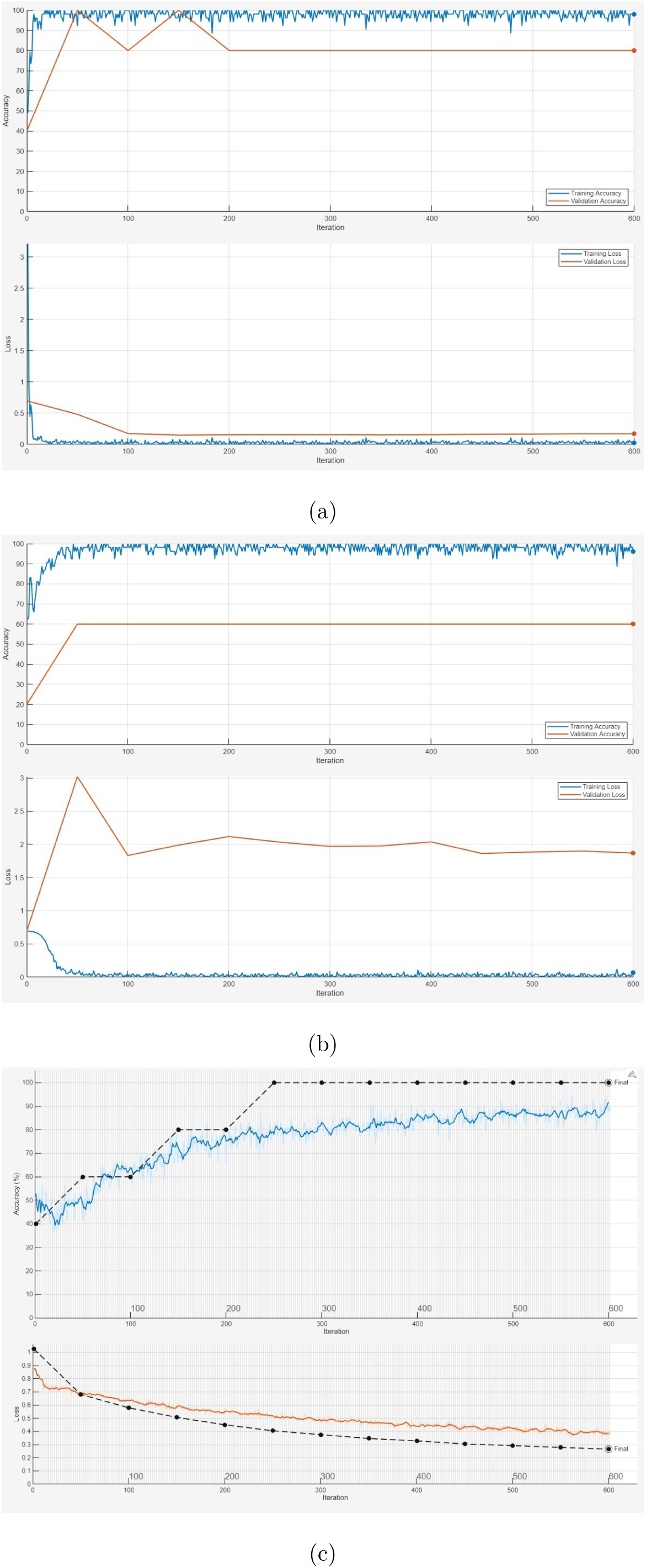
Training and validation processes: 1D-CNN (a), LSTM (b), and BERT feature based net, where solid and dotted lines indicate training and validation, respectively (c).

**Figure 5:**
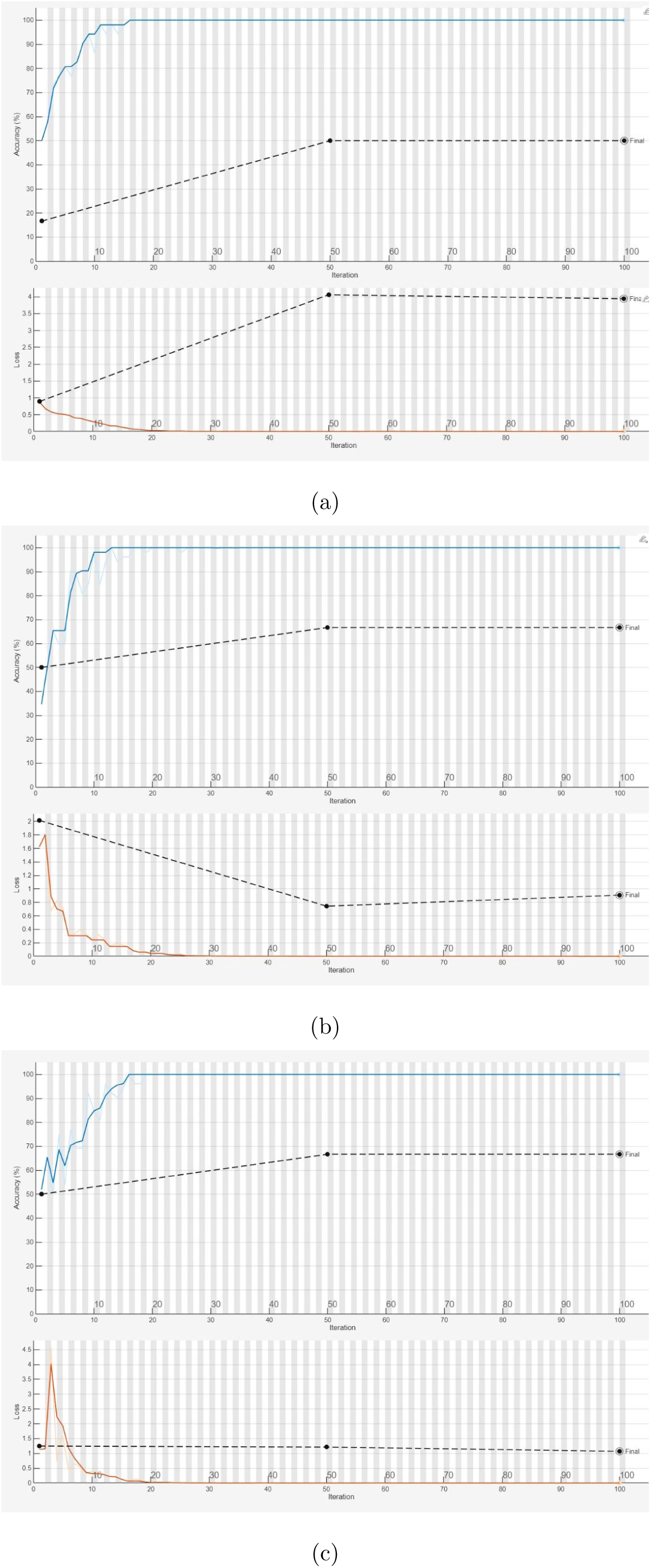
Training and validation processes of pretrained CNN models: SqueezeNet (a), GoogLeNet (b), and AlexNet (c), where solid and dotted lines indicate training and validation, respectively.

**Table 3:** Average performance measures of AI models.

| Model | ACC (%) | SEN (%) | SPE (%) | PRE (%) | F1 | AUC |
| --- | --- | --- | --- | --- | --- | --- |
| SqueezeNet | 50.00 | 46.67 | 53.33 | 54.17 | 0.49 | 0.62 |
| GoogLeNet | 56.67 | 66.67 | 46.67 | 58.67 | 0.61 | 0.62 |
| AlexNet | 66.67 | 73.33 | 60.00 | 63.33 | 0.67 | 0.73 |
| LSTM | 56.67 | 75.00 | 41.67 | 55.56 | 0.62 | 0.72 |
| BERT | 76.67 | 83.33 | 66.67 | 83.33 | 0.82 | 0.73 |
| 1D-CNN | 84.00 | 86.67 | 86.67 | 86.67 | 0.84 | 0.93 |

## 4 Discussion

### 4.1 The LSTM model

This classifier showed a sensitivity of 75.00%, indicating it correctly identified periapical infections in three out of every four instances. However, its specificity is much lower at 41.67%, suggesting a significant struggle in accurately classifying caries. This imbalance between sensitivity and specificity highlights that the LSTM model was biased toward recognizing periapical infections but failed to detect caries effectively. The overall performance is further reflected in the model’s moderate precision of 55.56%, F1 score of 0.62, and a relatively low AUC of 0.72, indicating that while it performed reasonably well in identifying caries, its ability to distinguish between the two classes was limited.

The low performance of the LSTM model, particularly in terms of specificity when classifying caries, can be attributed to several factors. First, the LSTM’s ability to capture sequential dependencies in text might be limited by the small dataset size. The model may not have enough data to learn the subtle differences between caries and periapical infections from the text alone, leading to a bias toward identifying one class (such as periapical infections) more accurately than the other. Moreover, LSTM models are typically better suited for sequential data such as time series, but they may not be the most effective for text classification tasks without appropriate preprocessing or feature engineering.

### 4.2 The BERT model

BERT achieved a sensitivity of 83.33%, indicating strong performance in correctly identifying periapical infection cases. However, its specificity was lower at 66.67%, suggesting a relatively higher misclassification rate for caries. While the model effectively detected periapical infections, it struggled to distinguish periapical infections from caries, which is reflected in its overall accuracy of 76.67%. The precision of 83.33% indicates that most of the positive classifications were correct, contributing to a high F1 score of 0.82, which balances precision and sensitivity. However, the AUC of 0.73 suggests moderate overall discriminative ability. This performance could stem from challenges in capturing subtle textual differences in descriptions of caries.

BERT’s moderate performance may be due to reliance on textual descriptions, which can be ambiguous or inconsistent. The small dataset limits learning diverse patterns, increasing the risk of overfitting to specific linguistic variations. Additionally, medical image interpretation requires nuanced domain knowledge that text alone may not fully capture. Unlike image-based models that analyze spatial and structural features, BERT depends solely on textual abstraction, which may omit subtle visual details crucial for distinguishing between similar conditions like caries and periapical infections.

### 4.3 The 1D-CNN model

This net, on the other hand, demonstrated a much more balanced performance with both sensitivity and specificity at 86.67%. This indicates that the model performed well in identifying both caries and periapical infections with equal accuracy. The balance in performance is also evident in the model’s high accuracy of 84.00%, precision of 86.67%, and F1 score of 0.84. Additionally, with an AUC of 0.93, the 1D-CNN model exhibited strong overall classification capability, effectively distinguishing between the two classes with a high degree of robustness. While the 1D-CNN model demonstrated strong performance with balanced sensitivity and specificity, caution is also required when interpreting its results on the small dataset. Despite its high accuracy, the model may still face challenges in generalizing to larger or more diverse datasets. With a limited sample size, there is a risk of overfitting, where the model might learn patterns specific to the small dataset that do not hold well in broader contexts. The 1D-CNN’s performance should be validated on larger datasets to ensure its robustness and reliability in real-world applications.

### 4.4 Comparisons

As shown in Table 3, the classification performance of Group 1, which includes three pretrained CNN models trained on panoramic radiographs (SqueezeNet, GoogLeNet, and AlexNet), is generally lower than that of Group 2, which consists of three text-based classifiers (LSTM, BERT, and 1D-CNN).

Among the pretrained CNN models, AlexNet achieved the highest accuracy at 66.67%, with a sensitivity of 73.33% and a specificity of 60.00%, suggesting it performed better in detecting both caries and periapical infections compared to SqueezeNet and GoogLeNet. While GoogLeNet showed relatively balanced sensitivity and specificity, it struggled with an overall accuracy of 56.67%. SqueezeNet exhibited the weakest performance, with an accuracy of only 50.00% and poor sensitivity (46.67%). The AUC values for all the pretrained CNN models remained relatively low, ranging from 0.62 to 0.73, indicating moderate classification ability.

In contrast, text-based classifiers demonstrated superior performance. The 1D-CNN model outperformed all others, achieving the highest accuracy of 84.00% with balanced sensitivity and specificity at 86.67%, along with the highest AUC of 0.93. BERT also showed strong results, with an accuracy of 76.67%, though its specificity (66.67%) was lower than its sensitivity (83.33%), suggesting a bias toward detecting caries more effectively. The LSTM model struggled, with an accuracy of 56.67%, high sensitivity (75.00%), but very low specificity (41.67%), indicating difficulty in correctly identifying periapical infections.

Overall, text-based classification outperformed image-based classification in distinguishing caries and periapical infections from panoramic radiographs, particularly with 1D-CNN and BERT achieving significantly better results than the CNN-based models. The superior performance of text-based classifiers suggests that transforming radiographic data into textual descriptions enables more effective feature extraction, reducing dependence on large annotated image datasets and complex preprocessing.

### 4.5 Training and validation

Analysis of the training and validation processes of the 1D-CNN, LSTM, and BERT models using a hold-out data partition, as shown in Figure 4, can provide insights into the classification performance of each model.

For the 1D-CNN model (Figure 4(a)), the training process appeared stable, with both training and validation performance improving steadily. The small gap between the solid (training) and dotted (validation) lines suggested minimal overfitting, indicating that the model generalized well to unseen data.

In the case of the LSTM model (Figure 4(b)), the training curve improved, but the validation curve may show more fluctuations or a larger gap, suggesting potential overfitting or difficulty in generalizing. LSTMs often struggle with smaller datasets, as they require longer sequences to fully leverage their sequential learning capabilities.

For the BERT-based model (Figure 4(c)), validation accuracy reached 100% while validation loss decreased, suggesting strong learning. However, this raises concerns about overfitting, as the small dataset may have led the model to memorize patterns rather than generalize. Further evaluation on independent data or a larger dataset is needed to confirm its reliability and real-world applicability.

The training and validation processes of SqueezeNet, GoogLeNet, and AlexNet exhibit overfitting, as illustrated in Figure 5. All three models reached 100% training accuracy while maintaining much lower validation accuracy. SqueezeNet struggled the most, with unstable validation accuracy and erratic validation loss, indicating poor generalization. GoogLeNet showed better training stability, but its validation accuracy fluctuated significantly, suggesting that the model memorized training patterns but failed to generalize effectively. AlexNet performed relatively better, with a more consistent validation accuracy trend, though still affected by overfitting. The high validation loss reinforced the limited ability of these pretrained CNNs to classify dental diseases from panoramic radiographs reliably.

### 4.6 Image-based text descriptions

Using ChatGPT to translate panoramic radiographs into text descriptions offers several advantages over direct classification of the images using image-based classifiers as follows.

#### Ability to Leverage Semantic Understanding

ChatGPT can analyze and describe high-level features of radiographs in natural language, summarizing complex patterns or abnormalities in a human-readable format. These textual descriptions can encapsulate contextual information and insights that may help in downstream classification tasks.

#### Simplified Workflow

Generating text descriptions directly from radiographs can simplify the workflow by reducing the preprocessing steps needed for image-based classifiers. This allows for faster and more efficient analysis, particularly in large-scale datasets.

#### Scalability and Resource Efficiency

Reducing image resolution (by 25 times) simplifies the computational load and storage requirements, which is particularly useful for large-scale datasets. Image-based classifiers often struggle with reduced-resolution images, as essential visual details may be lost. In contrast, ChatGPT can still infer potential abnormalities from simplified features, potentially acting as a bridge between low-quality inputs and meaningful outputs.

#### Improved Generalizability

Instead of relying solely on pixel-level patterns, textual descriptions generated by ChatGPT can generalize better across variations in image quality, acquisition settings, or equipment. This approach can reduce the dependency on extensive image-based model training, which often requires high-resolution data and large labeled datasets.

#### Enhanced Interpretability

Text-based outputs are inherently more interpretable to healthcare professionals than direct image classifications, making it easier to validate the results and integrate them into diagnostic workflows. Descriptions can highlight specific features, such as “possible bone loss near tooth x”, rather than providing a simple categorical label.

## 5 Conclusions

This study demonstrates the potential of using a large language model to translate panoramic radiographs into textual descriptions for dental disease classification using deep learning. By leveraging text-based classification instead of direct image-based analysis, the approach eliminates the need for complex image preprocessing, such as segmentation, and allows models like 1D-CNN to process text-based descriptions efficiently. Compared to traditional image-based classifiers, this method offers greater interpretability and accessibility, particularly in settings where high-resolution imaging or expert annotations are limited.

Beyond dental disease classification, this approach has broader applications in medical imaging, where textual descriptions can be used for AI-driven diagnosis across radiology, dermatology, and pathology. Future research should focus on optimizing text generation from images, ensuring that descriptions capture clinically relevant features with high fidelity. Additionally, investigating hybrid models that combine text-based and image-based features may enhance classification accuracy and generalizability. Expanding datasets, improving domain-specific language models, and incorporating external clinical knowledge into text-based AI models will be crucial for advancing AI-assisted diagnostics in healthcare.

## Author contribution

TDP contributed to conception, technical design, computer coding and implementation, data interpretation, and writing the manuscript.

## Data availability

ChatGPT-generated text data file is freely available at the author’s personal website: https://sites.google.com/view/tuan-d-pham/codes, under the title “Image-to-text classification of dental diseases”.

## Software availability

MATLAB codes implemented in this study are freely available at the author’s personal website: https://sites.google.com/view/tuan-d-pham/codes, under the title “Image-to-text classification of dental diseases”.

